# COVID-19 NFL Injury Prevalence Analysis, A Follow-Up Study

**DOI:** 10.1101/2023.01.11.23284292

**Authors:** Troy B Puga, Josh Schafer, Grace Thiel, Nicholas Scigliano, Tiffany Ruan, Andres Toledo, Prince N Agbedanu, Kevin Treffer

## Abstract

**Background:** In 2020, COVID-19 spread across the world and brought the world to a halt, causing the shutdown of nearly everything in order to prevent its spread. The NFL experienced the same effects of the shutdowns leaving athletes unable to train in some of the most advanced facilities with many of the best trainers in the world. Through a previous study, *COVID-19 Return to Sport Injury Prevalence Analysis*, it was determined that there was increased injury prevalence during the 2020 season likely due to decreased physiological adaptations within athletes bodies that resulted from facility shutdowns. Understanding injury epidemiology is vital in the prevention of injuries and the development of return-to-play protocols.

**Objective:** The objective of this study is to perform a follow up study to *COVID-19 Return to Sport Injury Prevalence Analysis* in order to to examine the longitudinal effects of the COVID-19 pandemic on injury epidemiology. This study will examine if there was a recovery to baseline or lingering effects from the COVID-19 pandemic-induced spike in injuries.

**Methods:** Injury tallies collected from the 17-week-long 2020 NFL regular season, played after COVID-19 restrictions, were compared with the injury tallies collected from the 18-week-long NFL regular seasons (2021, 2022), in order to determine if there was a change in injury prevalence. An unpaired t-test was conducted to compare the mean injuries per team per week between each of the 2020, 2021, and 2022 regular seasons.

**Results:** The 2022 and 2021 NFL regular seasons produced lower numbers of total injuries than the 2020 NFL regular season that was impacted by COVID-19. The comparison of the mean number of injuries per team per week of the 2020 season compared with the 2021 regular season was statistically significant (*P*=.03). The comparison of the 2020 and 2022 regular seasons was also statistically significant (*P=*.02).

**Conclusions:** The results of this follow-up study and our previous study show that extended training interruptions have the ability to induce detraining and lead to increased injuries.

Additionally, the results of this study show that retraining can occur and lead to injury protective factors. This is the first large scale opportunity to demonstrate the effects of these principles and how they are important to understanding injury epidemiology.

## Introduction

In 2020, the COVID-19 pandemic spread across the world and led to the shutdown of everything but essential services. Sports were no exception, as competition came to an immediate halt. The National Football League (NFL) was one such sport that faced a shutdown, as they closed all facilities from late March 2020 into late May 2020 [1,2]. A previous study conducted entitled *COVID-19 Return to Sport: NFL Injury Prevalence Analysis* examined the effects of this shutdown on injury epidemiology [2] This study showed that there was an increased injury prevalence during the 2020 NFL season which was impacted by the COVID-19 pandemic [2]. The study concluded that the increased injury prevalence was due to the decline in athlete training as a result of the shutdowns [2].The decline in training, hence, the lack of bodily preparation for the 2020 NFL Season resulted in decreased physiological of their bodies to strenuous exertions, which are a hallmark of the sports [2]. This is a follow-up study to examine the longitudinal effects of the COVID-19 pandemic shutdowns on injury prevalence in the NFL following the 2020 NFL season.

The NFL is an American Football league composed of 32 teams with some of the best athletes in the world. The NFL is widely recognized as having some of the best trainers and facilities to ensure that these high level athletes are prepared to play the season. NFL athletes follow intense training in order to physically prepare their bodies for the demand of a strenuous season [3]. While training may not prevent every injury, research has shown that training produces a protective effect from injury [4-6]. Traditionally, the NFL played a 17-week NFL season yearly until the 2021 season, when they transitioned to an 18-week long NFL season for all future seasons [7].

Our previous research demonstrated that the COVID-19 pandemic had acute effects on injury prevalence during the 2020 NFL season [2]. Much of this can be attributed to the decreased physiological adaptations that occurred during shutdowns, when athletes were unable to access NFL training facilities [1,2]. These athletes most likely exhibited effects from detraining, a process in which athletes undergo a loss of physiological and performance-based adaptations from previous training due to a lack of sufficient training [8,9]. It is believed that many athletes across the US and the world most likely exhibited effects of acute detraining during the COVID-19 shutdown [9,10]. Previous shutdowns in the NFL have shown that detraining can occur, such as during the 2011 lockout where the preseason saw increased achilles tendon injuries [11]. Training is one of the most important interventions for injury prevention and improved athletic performance in athletes of all populations [4-6,12-14].

The effects of detraining are a very serious concern in regards to injury epidemiology. We can clearly see that detraining has significant potential to lead to injuries; however, complete recovery from detraining can be obtained through a sufficient training program [9,15]. The amount of time necessary to recover from the effects of detraining is widely debated [9]. The amount of time for training adaptations often differ between people based upon training levels, genetics, and a host of other factors [9,16-18]. Many of these genetic and environmental factors are considerations in the variability of injury recovery times, and why some athletes may recover more quickly [19-21]. While we know recovery is complex and difficult to predict, research has suggested that the time necessary to fully return from detraining is often similar or longer to the length of detraining [15]. The variations of training adaptations and recovery from detraining makes the examination of the longitudinal effects of the COVID-19 pandemic that much more important to understanding longitudinal injury epidemiology.

Longitudinal research on injury epidemiology due to a major event such as the COVID-19 pandemic is scarce. The goal of this research is to understand the effects of a major event such as the COVID-19 pandemic on longitudinal injury epidemiology. We hypothesize in this study that injury prevalence for the 2021 and 2022 seasons will be lower than the 2020 NFL season that was affected by the COVID-19 pandemic shutdowns. We hypothesize, in this study, that lower injury prevalence is due to athletes having full-access to sports performance facilities and staff to properly prepare their bodies during the 2021 and 2022 seasons. It is believed during these two seasons, athletes had adequate time and preparation to induce the necessary physiological changes and undergo retraining to avoid the effects of detraining observed following the shutdowns of the COVID-19 pandemic.

## Methods

### Study Design

The methodology for this study design was adapted from our previous study *COVID-19 Return to Sport: NFL injury Prevalence Analysis* [2]. Data for the 2020 NFL regular season was collected in our previous study and used for further analysis [2]. The number of injuries was tallied for the 18-week (2021, 2022) NFL regular seasons using the weekly published injury reports by each NFL team. Injury reports are made publicly available by each team in the NFL. If an official injury report was not available through the individual team media, a deferment was made to the official NFL website. Athletes listed with the same injury for consecutive weeks were only counted once, unless they presented with an injury to a different anatomical region. Contact injuries were included in this study as this is a non-modifiable risk factor that can not be controlled due to football being a contact sport [2,22]. COVID-19, sick days, and non-medical days off were not included in the injury tally. Illnesses were not included in this study for the fact that illnesses are not considered a physical injury, and should be reported separately from injuries when performing injury epidemiological studies [2,23]. The 2020 season, which was played after COVID-19 shutdowns, was used for comparison to the subsequent 2021 and 2022 seasons. Since the NFL regular season in 2020 was 17 weeks, and the 2021 and 2022 regular seasons were changed to 18 weeks, analysis was conducted using the mean number of injuries per team per week. This adjustment allowed for comparison of two different regular season lengths.

### Data Analysis

A data analysis was performed on the collected data. The data analysis for this study was changed from the mean number of injuries per team from the previous study [2] to the mean number of injuries per team per week to correct for the change of the 17-week NFL season to the 18-week NFL season in 2021 and 2022 [7]. The 2020 regular season mean number of injuries per team per week was compared to the 2021 regular season mean number of injuries per team per week using an unpaired t-test in a similar fashion to our previous study [2]. The 2020 regular season mean number of injuries per team per week was compared to the 2022 regular season mean number of injuries per team per week using an unpaired t-test. The 2021 regular mean number of injuries per team per week and 2022 regular season mean number of injuries per team per week were also compared using an unpaired t-test.

**Figure 1:**
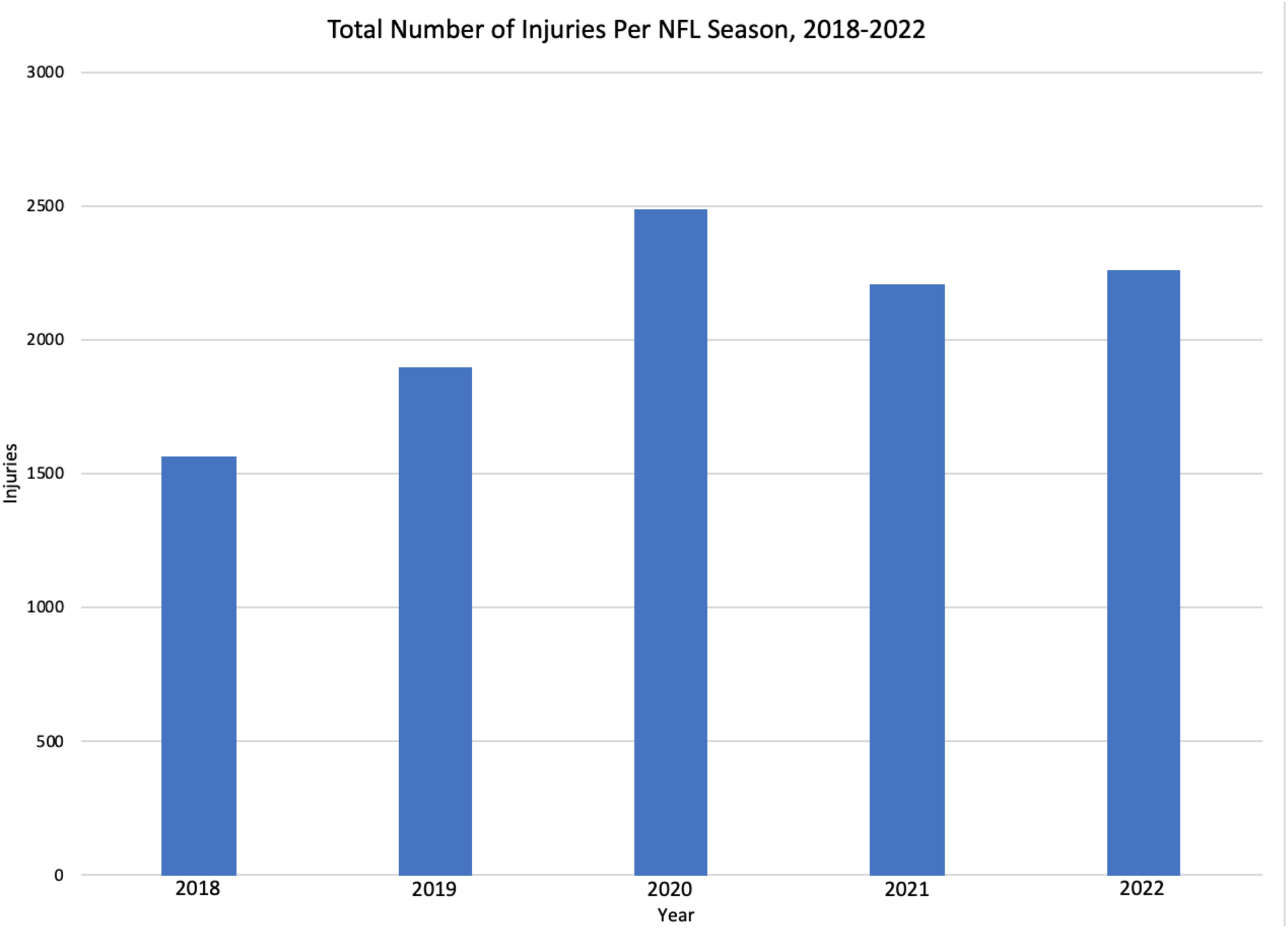
Total number of injuries per NFL regular season including the 2018, 2019, and 2020 17-week NFL seasons from our previous study [2], and the 2021 and 2022 18-week NFL seasons.

**Figure 2:**
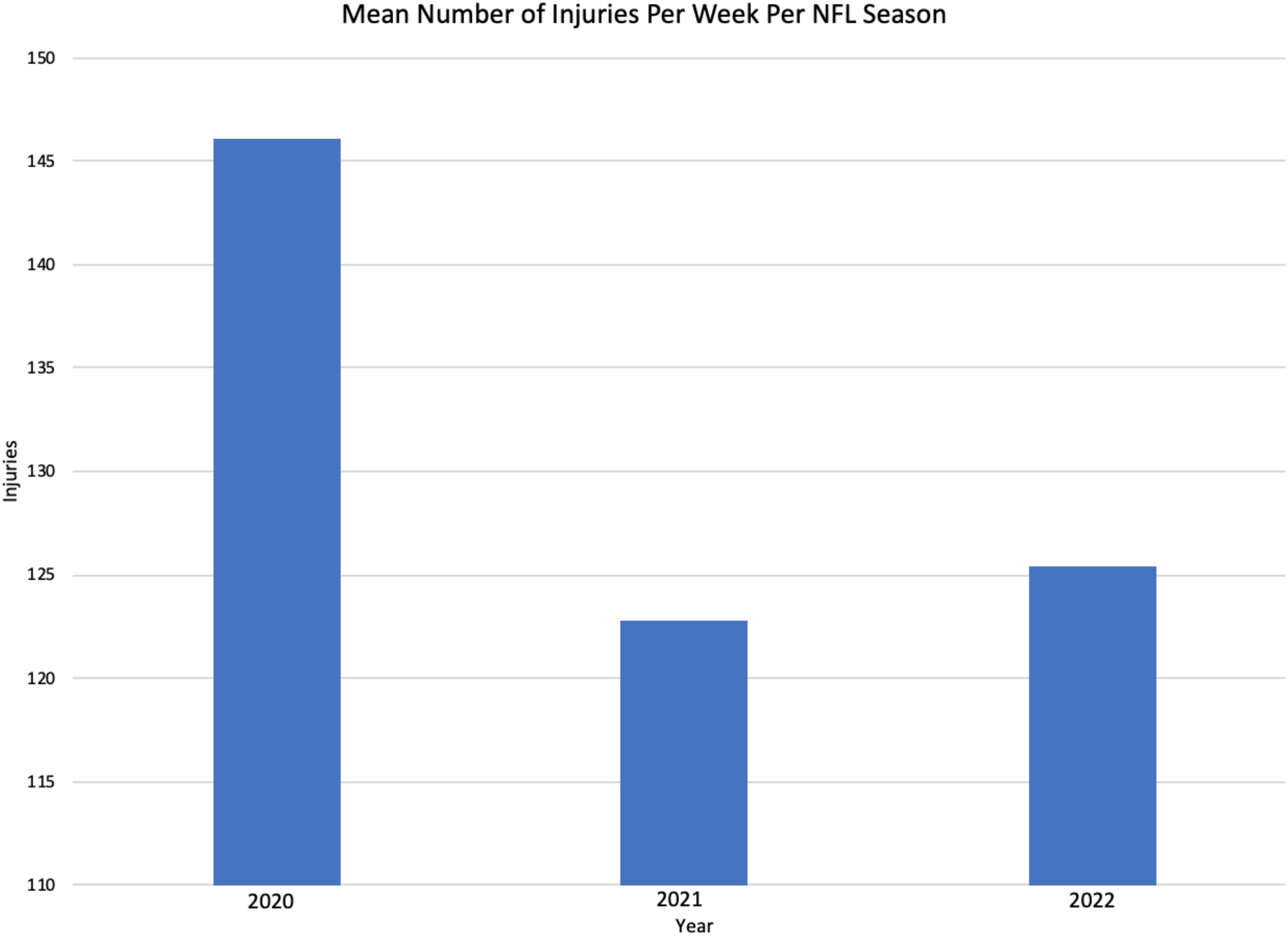
League wide mean number of injuries per week per NFL regular season for the 2020, 2021 and 2022 seasons.

## Results

The injury tally for the 2020 season was collected in our previous study [2]. The 2020 season produced a total of 2484 injuries in our previous study [2]. This number was divided by the 17, the number of weeks in the NFL regular season for 2020, to allow appropriate comparison with the change to an 18-week season adaptation in 2021 and 2022. In this further analysis, the 2020 season produced a league-wide mean number of injuries per week of 146.1, and a mean number of injuries per team per week of 4.6. The 2021 season produced a total of 2210 injuries, and was divided by 18, the number of weeks in the 2021 regular season. This produced a league wide mean number of injuries per week of 122.8, and a mean number of injuries per team per week of 3.8. The 2022 season produced a total of 2257 injuries, and was divided by 18, the number of weeks in the 2022 regular season. This resulted in a league wide mean number of injuries per week of 125.4, and a mean number of injuries per team per week of 3.3.

Data analysis with an unpaired t-test found that the mean number of injuries per team per week for the 2020 COVID-19 NFL season when compared to the mean number of injuries per team per week during the 2021 season produced a statistically significant difference *(P*=.02). The unpaired t-test comparison of the mean number of injuries per team per week between the 2020 and 2022 seasons produced a statistically significant difference as well (*P*=.03). Comparison of the mean number of injuries per team per week of the 2021 and 2022 seasons did not produce a statistically significant difference (*P*=.79).

## Discussion

Our study results showed statistically significant differences in comparison of the 2020 COVID-19 impacted NFL season to the subsequent 2021 (*P*=.02) and 2022 (*P*=.03) NFL seasons. The study also showed that there was a lower total number of injuries across the league in 2021 (2210 injuries) and 2022 (2257 injuries) when compared with the 2020 (2484 injuries) COVID-19 impacted NFL season. This occurred even with the addition of an extra week of the regular season in the 2021 and 2022 seasons when compared with the 2020 season [7]. Our previous study showed the impact of the COVID-19 pandemic, which led to a higher number of injuries in the 2020 NFL season when compared with earlier seasons due to decreased training adaptations stemming from lockdowns of facilities to help mitigate the spread of COVID-19 [2]. This study demonstrated recovery from that spike back to pre-pandemic season levels.

Previous research has demonstrated the effects of shutdowns and detraining, which lead to a negative impact on athletic performance and health [2,8-11]. Preparation for athletic competition is vital for the health and performance of athletes [4-6,12-14]. This follow-up study aimed to examine if the COVID-19 pandemic effects on injury epidemiology persisted, or if athletes were able to return their bodies to peak pre-pandemic shape. The decline in injuries after the COVID-19 pandemic season in this follow-up study showed that athletes were able to return to pre-pandemic athletic preparation. This was most likely achievable through having a full off-season of full-access training for the 2021 and 2022 seasons. NFL players train in some of the best facilities in the world while under the supervision of renowned sports medicine personnel. We can conclude from this follow-up study that the COVID-19 pandemic did have an acute effect on injuries with a spike in the 2020 season. We can also deduce from this follow-up study that the COVID-19 pandemic did not have a longitudinal effect on injuries due to the decrease in injuries during the 2021 and 2022 regular seasons when compared to 2020. This follow-up study further suggests that athletic preparation through performance training is vital for the preparation of sport and preventing injury.

We do believe our study was not without limitations, and these limitations are consistent with our previous study. Similar to our previous study, we believe that our study was limited by the potential of underreporting of injuries by players [2]. Another potential limitation, consistent with our previous study is that we did not know the exact difference in training hours [2]. However, we know that there were limitations for training in 2020 [1], and believe that survey would be limited due to recall bias [2]. We believe that despite these limitations, this study provides an accurate representation of the effects of detraining on injury epidemiology.

This research has demonstrated the importance of performance training for sport on injury epidemiology. This study shows that recovery from detraining is possible under the proper training conditions, in which athletes will induce adaptation and preparation for sport. This is the first large scale opportunity that has been available to study detraining and retraining principles. Findings from this study serve as a foundational piece of research in proving what many have hypothesized in regards to training and injury prevention.

Further studies must determine exactly how much time is needed to return to sport from a major event such as the COVID-19 pandemic, injuries, or time off. Qualified performance coaches and rehabilitation professionals are vital for helping to solve these issues. Further research must also be done at the collegiate and amateur level to examine the effects of detraining and retraining, as these individuals may not have the access to the care and facilities that NFL players receive. The research from the previous study and this follow-up study is a start to the future discussion of necessary preparation times and preperation levels. Further injury epidemiological trials and observational studies must be conducted to continue the evolution of return to sport protocols and training programs to combat detraining in adverse situations.

## Data Availability

All data produced in the study is available online through the NFL and team websites.

## Author Contributions

All authors contributed equally to this research.

## Ethical Approval

This study did not require institutional review board approval as all data is publicly available and does not qualify as human subjects research per United States Department of Health and Human Services policy 45 CFR 46.102.f

## Abbreviations

NFL: National Football League

